# Utilization of Family Planning and Associated Factors among Reproductive Age Women with Disability in Arba Minch Town, Southern Ethiopia

**DOI:** 10.1101/19013821

**Authors:** Yibeltal Mesfin Yesgat, Feleke G/Meskel, Wubshet Estifanous, Yordanos Gizachew, Seid Jemal, Natneal Atnafu, Keyirden Nuriye

## Abstract

**Introduction:** globally, one type of contraceptive was used by around 63 percent of women. Women with disabilities account for 10 percent of all women and make up three-quarters of the disabled people in low and middle-income countries.

**Objective:** to assess utilization of family planning and associated factors among reproductive-age women with disability group in Arba Minch town, southern Ethiopia from 1^st^ March to April 15, 2019.

**Methods:** community-based cross-sectional study with simple random sampling was used to select 4l8 reproductive age women with disabilities. Data were collected using a structured questionnaire and interview by eight trained females who completed grade twelve two of which communicate by speaking and sign language. Data were entered using Epi info 7 and exported to SPSS version 20 for analysis. A statically significant variable in the final model was declared by AOR, 95%CI and p-value <0.05.

**Result:** in the current study family planning utilization among all reproductive-age women with disabilities was 33.7%. Family planning utilization was 2.2 times higher among those who have employed compared with those with not employed (AOR2.2 95% CI, 1.77-4.15). Women who had a positive attitude were 2.3 times more likely to use family planning than negative attitudes (AOR 2.3:95% CI, 1.21- 3.87). Besides these women who got married were almost four times more likely to use family planning methods than unmarried (AOR 3.9:95% CI, 2.31-6.63).

**Conclusion:** The level of family planning utilization was low among reproductive women with disabilities and factors associated were attitude, marital status, & employed status, therefore governmental and non-governmental organization should promote for women with disabilities to change the attitude and creating job opportunities.

## Introduction

Family planning allows individuals and couples to anticipate and attain their desired number of children and the spacing and timing of their births. It is achieved through the use of contraceptive methods and the treatment of involuntary infertility (1).

Access to and uptake of family planning is important directly or indirectly, for achieving the recently launched Sustainable Development Goals (SDGs). It can contribute to reducing chronic poverty and hunger, increasing access to quality education, Address the Challenges of Climate Change, and ensure sustainable consumption, enhancing gender equality, improving and maternal health and reducing childhood mortality through its role in preventing unwanted pregnancy and reducing fertility rate as a whole (2, 3)

Family planning is used by the majority of married or in-union women in almost all parts of the world. Worldwide in 2017, 63 percent of married or in-union women of reproductive age were using some form of contraception, including any modern or traditional methods of contraception. From this woman with disabilities comprise 10 percent of all women and make up three-quarters of the disabled people in low and middle-income countries (4, 5)

In Africa, 36% of reproductive-age women use any form of contraceptive but still 22 % to unmet need for modern contraception of this the major proportion is shared by Sub-Saharan Africa which is highest in the world(6, 7).

Even those reports of 2016 Ethiopia demographic health survey (EDHS) don’t report contraceptive use of disabilities women separately 36 percent of currently married women and 58 % unmarried sexually active women use any form of contraceptive (8).

Family planning among disabled is neglected but disabled have the same need for reproductive health care as a general population. Persons with disabilities face many barriers to care and information about SRH. The most frequent assumption that persons with disabilities are not sexually active and do not needs SRH service (11, 12).

Factors associated with failure to meet need family planning for disabled women vary from country to country when compared with non-disabled women. According to World Health Organization (WHO) the highlighted reasons are: limited choice of family planning methods, limited access to contraception, poorness, fear or experience of having side-effects, cultural or religious opposition, poor quality of available family planning services and gender-based barriers pregnancy, concern of health problems, lack of disability-related clinical services, and stigma and discrimination(13).

In many sub-Saharan African(SSA) countries including Ethiopia factors affecting use contraceptive are situations like level of education, desire more children, age, side effect of contraception, transportation problems, lack mass media avail for disable and husband and communities attitude barriers to health services include: lack of physical access, including transportation and/or proximity to clinics and, within clinics, lack of ramps, adapted examination tables, and the like; lack of information and communication materials (e.g. lack of materials in Braille, large print, simple language, and pictures; lack of sign language interpreter)(14-16).

Finally, in our country, there were a lot of studies that were conducted to assess the magnitude of family use and associated factors among reproductive-age women in general but few kinds of research only addressed women with disabilities. Specifically, research conducted previously are institution based which focus on women with disabilities supported by association with the sexual and reproductive health (SRH) service facility for disabled avail and also those article miss some important factors like attitude. Therefore, this study aimed to assess the utilization of family planning and the factor associated with women with disabilities in Arba Minch town.

## METHODS AND MATERIALS

### Study design and setting

A community-based cross-sectional study was conducted from 1st March to 15, April, 2019Arba Minch town is found in the Gamo zone, the Southern Nations, Nationalities and Peoples Region. It is located at 505km distance south of Addis Ababa (the capital city of Ethiopia) and 275km southwest of Hawassa (capital town of the regional state). Based on 2007 census Arba Minch town has a population of 74,879 of which 39,208 (52.36%) are men and 35,671 (47.64%) are women from this the proportion of people living with disabilities in Arba Minch town was about 638 and increased to 1561 in 2018 from these women in reproductive age occupied 462.

### Study population

Reproductive age women with disabilities group live in Arba Minch town known by Arba Minch town labors and social affairs

### Sample size determination

The minimum sample size was calculated by using a single population proportion formula with P = 44.4 %(proportion of utilization of family planning in Addis Ababa city).Level of significance 5% (α=0.05), the margin of error 5% (d=0.05), and 10% non-responses rate Thus, a total sample of 418 women with reproductive age with the disability group participated.

### Sampling procedure

Based on the report of the Arba Minch town administrative labor and social affairs disabled person female gender was selected and registered with an excel sheet by their identification card (ID) and age. Sampling frames were developed by selecting 462 reproductive age women with disabilities to form a list obtained from the Arba Minch town administration, labor and social affairs with their age. Finally with the help of their ID numbers of each 418 women select by lottery methods

### Data collection tool

Data was collected by adopting Ethiopian Demographic and Health Survey. Semi structured questionnaire was first prepared in English and then translated into the Amharic language then back to English for checking language consistency(8). The main components of the questionnaire are socio-demographic characteristics, knowledge of the respondents towards family planning methods, attitude, and reproductive health-related factor and client-related factors.

### Data collection procedures

Data was gathered using eight grade twelve complete. Two of the collectors were able to communicate by sign language to obtain information from the deaf. The data collectors were trained for two days before the actual data collection on interview strategy and data recording. Lecturing, mock interview and actual field practice were used to train data collectors. The data collection was done under close supervision

The interview was used for data collection. To collect data from the deaf the two females who can speak sign language were used and for the blind since the questionnaire was interviewed their blindness was not affecting the process.

### Data processing and analysis

All filed questioners were cheeked for completeness, consistency, and accuracy, then the data were entered into Epi info version 3.7 then exported to statically package of social science (SPSS) version 20 for data analysis.

Bivariate analysis with a crude OR (COR) of 95% CI was used to assess the degree of association between each independent variable and the outcome variable by using binary logistic regression. Independent variables with a P-value of ≤0.25 were included in the multivariable analysis to control confounding factors. Multicollinearity was checked to see the linear correlation among the independent variables by using the variance inflation factor (VIF) and standard error (SE). Variables with a VIF of >10 and a SE of >2 were dropped from the multivariable model. The fitness of the model was checked by Hosmer-Lemeshow goodness of fit test not statically significant p-value.0.679. Crude and adjusted odds ratios along with 95% of confidence interval were used to identify factors associated with the outcome variable. The level of statistical significance was declared at a p-value of less than 0.05.

## Result

### Socio-demographic characteristics of the respondents

A total of three hundred ninety-eight reproductive age women with disability groups have participated with a response rate of 95.2%. The age of women ranges from 15 - 40, with the mean age of 23.15 (SD ± 5.1) years. About ninety-six women (24.1%) of women and twenty-four (15.1%) of partners of those who are married/union never been enrolled in formal education. A total of one hundred twenty respondents were employed represented by (31.2%) while 118(29.6%) was a student and 107(26.9%) were unemployed. Respondents who were protestant followers accounted for 288(72.4%), followed by 99(24.9%) who were orthodox and the rest were others including Muslim, Catholic, Adventist 11(2.9%). About 76(19.1%) of the respondents were categorized as having low household income and 86(21.6%) as moderate while 236(59.3%) were categorized as having high household income **(table 1)**.

### Reproductive health characteristics of the study population

A total of two hundred forty women of those has started sexual intercourse. Eighty-five respondents have given birth of which only 11.76% of the respondents had experienced child loss or death. A majority (63%) of the women reported having the desire to have children after two years and 8.8% desired no or no more children while the remaining 28.2% want within two years (**table2)**.

### Client characteristics related factors

One hundred forty women with disabilities discuss with their partner. On the other end, 69.2% supported by their partner to use family planning. Majority disabilities around 71 percent of the women are physically disabled.

One hundred eighty-seven reproductive age women with disabilities are exposed to media and the rest are not.

### General knowledge about family planning

The collected information regarding knowledge of contraceptive methods was after describing each method and asking respondents if she had heard of it. Using this approach, interviewers collected information about 9 modern family planning methods and two traditional methods. Knowledge of at least one method of contraception likely to have about it is 79.9%.

### Attitude towards family planning use

Concerning the level of attitude towards family planning, about 41 % of the respondents have negative Attitudes. More than half of the respondents (59%) had positive attitudes towards family planning use

### Facility related factor

Almost more than half of reproductive-age women with disabilities think SRH serves was not friendly.

### The proportion of family planning use

Around(one-third of the participants)134 (33.7 %:95% CI, 0.29-0.38)) of all reproductive-age women with disabilities are currently using a contraceptive. The most frequently used contraceptive methods are injectable 50% (n=67) followed by implants 25% (n=34).

### Reason for not using contraceptive

Study participants were asked why they are not currently using family planning. From the multiple reasons they have given the highest reason was being single with 134 frequencies.

### Factors affecting family planning utilization

Variables Age of the women, Marital status, women education, Media exposure, Occupation women, Household income, Attitude, Parity, knowledge and thinking sexual and health service friendly signify who are significantly associated with dependent variable at bivariate analysis and P-value <0.25 were further analyzed in the multivariate analysis to identify their related effects with family planning.

The odds of women to use family planning methods were AOR=2.2(1.77-4.15) times higher among those who have employed compared with those with not employed.

Women with disabilities who had were positive attitude AOR=2.4(1.43-3.98) time to use family planning methods compared with those who were a negative attitude

The study also indicated that the odds of family planning use were AOR=3.95 (2.33-6.70) times higher among those women with disabilities who had currently married on the use of family planning methods compared with those who did not have currently married**(table3)**.

## Discussion

This community-based research aimed to identify the use of family planning and associated factor in Arba Minch town among women of reproductive age with disability Finding from this study showed that the use of family planning was 33.7 percent and factors that contribute for use of family planning were occupation, marital status, and attitude.

In this study, the rate of usage for family planning was 33.7 percent (n=134) of all women of reproductive age with disabilities group. This outcome is greater than the research carried out in Bahirdar was 25.2 percent among 337 females of reproductive age with disabilities(10). This may be justified by increased access and inclusive sexual reproductive health services for the past four years.

In contrast, Addis Ababa’s study was greater than this finding, with contraceptive use being used by 44.4 percent of all women with disabilities in reproductive age(9). The reason for this difference may be the sample size and availability of health care facilities and disability-related family planning facilities in Addis Ababa.

This finding showed that the women with disabilities to use family planning methods were 2.2 times higher among those who have employed compared with those with not employed. This agrees with the Shembela refugee camp, Northern Ethiopia among reproductive-age women which favor being unemployed was 79 percent less likely to use contraceptive than employed(17). This suggesting that the ability of a woman to become employed empowered her to participate and take control of household decisions, including those regarding ideal family size and contraceptive use.

With regard to marital status, this study also indicated that the chances of family planning use among women of reproductive age with disabilities who presently married using family planning methods were almost four times greater than those who currently did not marry who endorsed by a study in the Oyo State, Nigeria of the married women were approximately four times more likely to use contraception than the single women(18).

To favor marital status study in the Dembia district, the use of married women in North West Ethiopia was 2.81 times more probable than unmarried females to use current contraceptives (19). The findings show the significance of couple inspiration in reproductive health problems including fertility and contraception through instruction and masculine involvement.

The other stronger factor for the grandstanded need for family planning was an attitude. Women with disabilities who had a positive attitude 2.4 time more likely to use family planning methods relative to those who had an negative attitude this finding almost similar to cross-sectional studies in the Nationalities of the Southern Nations and the People’s Region, Ethiopia among women of reproductive age group who state that positive attitudes towards contraceptives were almost twice than negative attitude (20).

However, this study is lower than study in Ghana and higher from Juba, South Sudan among reproductive-age women(21, 22). The possible reason for this difference may be socio-demographic character and the difference in the status of women’s habits of each country.

Finally, this study had some limitations since the data collection instrument addressed many questions concerning the use of contraceptives and sexual activity by interview for visual and hearing impairment respondent that it may be possible that they did not precisely respond. However, we try to fill the gap by using a female data collector.

## Conclusion

This finding showed around 33.7% of disabled women of reproductive age in Arba Minch town use any methods of family planning. Factors that are found to have associated with family planning use were women’s work status, marital status, and attitude. So we recommend that the Arba Minch town administration better enhance the employment status of women with disabilities. Persons with disability (PWD) Organizations, other governmental and NGOs should strengthen their family planning and SRH friendly service with special focus on unmarried women. The health extension workers and community nurses should increase their encouraged, educated women with disabilities to enhance a positive attitude for family planning.

## Data Availability

http://www.wku.edu.et/midwifery

## Declaration

## List of abbreviations

AOR: Adjusted odds ratio
EDHS: Ethiopian demographic health survey
CI: Confidence interval
COR: Crude odds ratio
FMOH: Federal ministry of health
SRH: sexual and reproductive health
PWD: the people with a disability

## Ethical clearance and consent to participate

Ethical clearance was obtained from an ethical clearance committee of Arba Minch University College of Medicine and Health Science. A permission letter was written to the Arba Minch town administration to conduct the study and another permission letter was also obtained from the Arba Minch town administration. Finally informed verbal and written consent was obtained from each respondent.

## Availability of data and materials

Full data set and other materials relating to this study can be obtained from the corresponding author upon reasonable request

## Author contributions

All authors contributed to data analysis, drafting or revising the article, gave final approval of the version to be published, and agree to be accountable for all aspects of the work.

## Acknowledgments

I would like to express my deepest appreciation Arba Minch University, College of Health Sciences, Department of Nursing for providing this opportunity. I am very grateful Arba Minch town labor and social affairs Administration and Arba Minch special need school staff for their great support. Finally, we would like to appreciate our data collectors, supervisors, questionnaire translators, and study participants; without them, the research would not be possible

## Tables and legends

Table 1 Frequency distribution of reproductive age women with disability by their background characteristics in Arba Minch town, 2019

Table 2 Distribution of reproductive age women with disability by reproductive health-related characteristics, Arba Minch town, 2019

Table 3 Factors associated with family planning, utilization among women with disability in Arba Minch town, 2019

